# Inhaled Prostacyclin Improves Oxygenation in Patients with COVID-19-induced Acute Respiratory Distress Syndrome

**DOI:** 10.1101/2021.11.15.21266343

**Authors:** Helene A. Haeberle, Stefanie Calov, Peter Martus, Lina Maria Serna Higuita, Michael Koeppen, Almuth Goll, Alexander Zarbock, Melanie Meersch, Raphael Weiss, Martin Mehrländer, Gernot Marx, Christian Putensen, Bernhard Nieswandt, Valbona Mirakaj, Peter Rosenberger

**Affiliations:** Department of Anesthesiology and Intensive Care Medicine, Tübingen University Hospital, Tübingen/Germany; Institute for Clinical Epidemiology and Applied Biometry, Faculty of Medicine, University of Tübingen, Tübingen, Germany; Department of Anesthesiology, Intensive Care and Pain Medicine, University of Münster, Münster, Germany; Department of Intensive Care Medicine, University Hospital RWTH Aachen, Aachen, Germany; Department of Anesthesiology and Intensive Care Medicine, University Hospital Bonn, Bonn, Germany; Institute of Experimental Biomedicine I, University Hospital Würzburg, Germany

**Author notes:** **<u>Address correspondence to:</u>** Peter Rosenberger, MD, PhD, Department of Anesthesiology and Intensive Care Medicine Universitätsklinikum Tübingen, Hoppe-Seyler-Straße 3, 72076 Tübingen, Tel.: +49 7071/29-86622. these authors contributed equally to this work (Valbona Mirakaj, Peter Rosenberger). **Author Contribution:** HAH, SV, MK, AG, BN, VM, PR – contributed to conceptualization and designed parts of the protocol; HAH, SV, MK, AZ, MM, RW, MaMe; GM, CP, VM – contributed significantly to acquisition of study data; PM, LMH – performed the power calculation, designed figures; VM, PR an PM – drafted the manuscript, collected approval by all authors.

**Keywords:** randomized clinical trial, Acute Respiratory Distress Syndrome, ARDS, COVID-19, sepsis, prostacyclin, oxygenation, inflammation

## Abstract

**Background:** Acute Respiratory Distress Syndrome (ARDS) results in significant hypoxia, and ARDS is the central pathology of COVID-19. Inhaled prostacyclin has been proposed as a therapy for ARDS, but data regarding its role in this syndrome are unavailable. Therefore, we investigated whether inhaled prostacyclin would affect the oxygenation and survival of patients suffering from ARDS.

**Methods:** We performed a prospective randomized controlled single-blind multicenter trial across Germany. The trial was conducted from March 2019 with final follow-up on 12^th^ of August 2021. Patients with moderate to severe ARDS were included and randomized to receive either inhaled prostacyclin (3 times/day for 5 days) or sodium chloride. The primary outcome was the oxygenation index in the intervention and control groups on Day 5 of therapy. Secondary outcomes were mortality, secondary organ failure, disease severity and adverse events.

**Findings:** Of 707 patients approached 150 patients were randomized to receive inhaled prostacyclin (n=73) or sodium chloride (n=77). Data from 144 patients were analyzed. The baseline oxygenation index did not differ between groups. The primary analysis of the study was negative, and prostacyclin improved oxygenation by 20 mmHg more than NaCl (p=0·17). Oxygenation was significantly improved in patients with ARDS who were COVID-19-positive (34 mmHg, p=0·04). Mortality did not differ between groups. Secondary organ failure and adverse events were similar in the intervention and control groups.

**Interpretation:** Although the primary result of our study was negative, our data suggest that inhaled prostacyclin might be a more beneficial treatment than standard care for patients with ARDS.

## Introduction

Acute respiratory distress syndrome (ARDS) is a common, life-threatening syndrome characterized by the development of severe hypoxia. The hallmark of SARS-CoV-2 infection is COVID-19-induced ARDS, which is associated with severe hypoxia. This hypoxia affects the function of secondary organs, and as a result, organ failure in the affected tissues may develop (1). The underlying cause of ARDS is uncontrolled and self-propagating inflammation within the alveolar space associated with the loss of pulmonary barrier function (2). Several pharmacological approaches have been tested in the past to improve oxygenation and overall outcomes of patients with ARDS with varying results (3-5).

Prostacyclins are used to treat patients with dyspnea due to pulmonary arterial hypertension, which is often associated with endothelial changes within the pulmonary vasculature (6, 7). ARDS, particularly COVID-19-induced ARDS, is characterized by pathological features such as endothelial injury, suggesting that prostacyclin therapy might be beneficial (8). A small, single-center observational study suggested that prostacyclins might improve oxygenation in patients suffering from ARDS. However, no systematic investigations have evaluated the effect of prostacyclin on a population suffering from ARDS (9). The aim of this trial was to test the hypothesis that prostacyclin would improve oxygenation and clinical outcomes of patients with ARDS, regardless of its cause (10).

## Methods

### Study design, Ethics and Oversight

We conducted a prospective randomized controlled, single-blind multicenter trial administering prostacyclin to critically ill patients with ARDS for 5 days. Two major changes in the design were amended in the protocol. First, patients who did not receive the study therapy according to the physician’s decision were included in the primary analysis population to avoid bias. Second, an extensive subgroup analysis was performed for patients with COVID-19, as the pandemic started during the study period. The study was approved by the Institutional Review Board of the Research Ethics Committee of the University of Tübingen (899/2018AMG1) and the corresponding ethical review boards of all participating centers. The trial was also approved by the Federal Institute for Drugs and Medical Devices (BfArM, EudraCT No. 2016-003168-37) and registered at clinicaltrials.gov (NCT03111212) For further details, please see Supplemental Data.

### Patients

Before the inclusion of patients into the study, the trial coordinators obtained consent for participation in the study. Only patients older than 18 years were allowed to enter the study. For details about inclusion and exclusion criteria please see Supplemental Data.

### Randomization and Interventions

Randomization was performed at a 1:1 ratio using a parallel group design. Randomization lists were generated at the biostatistical center using the software nQuery, release 4, and based on these lists, numbered envelopes were provided and used for randomization (stratified for center and using blocks of random length). For each center, a separate spate list was generated, and closed envelopes were supplied to the participating centers. Envelopes were opened only by the treating physician. The randomization number and treatment were recorded in the ID screening and enrollment list, dated and signed. The signed sheet was then stored at the participating center.

### Outcomes

The primary endpoint was the improvement in oxygenation defined as the oxygenation index on Day 5 of therapy. This outcome should not be affected by observation bias, as it is based on an objective routine measurement. Secondary outcomes included overall survival in the 90-day follow-up period; SOFA Organ Failure (SOFA) scores on Days 1-14, 28 and 90; duration of mechanical ventilation support; ICU length of stay; development of ventilator-associated pneumonia, pulmonary hemorrhage, gastrointestinal hemorrhage, pulmonary embolism, coagulopathy, delirium, ICU-acquired weakness and discharge location.

### Sample size

In a previous study of prostacyclin effect in 20 patients, an increase from 177±60mmHg to 213±67 mmHg was observed for PaO_2_/FiO_2,_ which was significant at the 0.01 level in an intraindividual comparison (9). Recalculation showed that the standard deviation was considerably smaller, as a p value of 0·01 corresponds to an effect size of 0·93 (intraindividual) and thus to an intraindividual standard deviation of approximately 40 in this study. For details about sample size see Supplemental Data.

### Statistical analysis

The primary hypothesis of the analysis was to show the superiority of inhaled prostacyclin to NaCl. The primary analysis population was the intention to treat the population of randomized patients and provide baseline values, except for six patients who were excluded for reasons documented in the Consort Flowchart. The primary endpoint, P_a_O_2_/FiO_2_, on Day 6 after baseline, i.e., Day 5 of prostacyclin treatment, was evaluated using a baseline-adjusted analysis of covariance model with the last measurement of p_a_O_2_/FiO_2_ before treatment serving as the baseline and the study arm and center as two-level factors. For further details see Supplemental Data.

## Results

### Enrollment and patients

The trial was conducted from March 2019 to August 2021. Seven hundred seven patients were screened for inclusion, of whom 150 patients were enrolled and randomized to receive either NaCl or prostacyclin (Iloprost®) inhalation 3 times/day for 5 days (Figure 1). The last patient was recruited on 14.05.21, and 144 patients were included in the primary analysis (n=72 NaCl, n=72 prostacyclin) since 6 patients withdrew consent during the course of the trial or during the observation period (n=4) or violated the inclusion criteria (n=2). The baseline characteristics of the patients are presented in Table 1. These characteristics were similar in both study groups (Table 1). The age of the intervention group was significantly higher than that of the control group at 61.5 years compared to 58.5 years. Regarding the pre-existing comorbidities, the group of patients treated with prostacyclin showed a higher incidence of pre-existing COPD and emphysema. The main causes of ARDS were COVID-19-induced ARDS, followed by bacterial infection that resulted in ARDS. Organ specific baseline characteristics and ventilation parameters did not differ between groups. There were more patients receiving extracorporeal membrane oxygenation (ECMO) therapy in the NaCl group than in the prostacyclin group (21 vs. 15), yet this difference was not significant (Supplemental Table 1).

**Figure 1.**
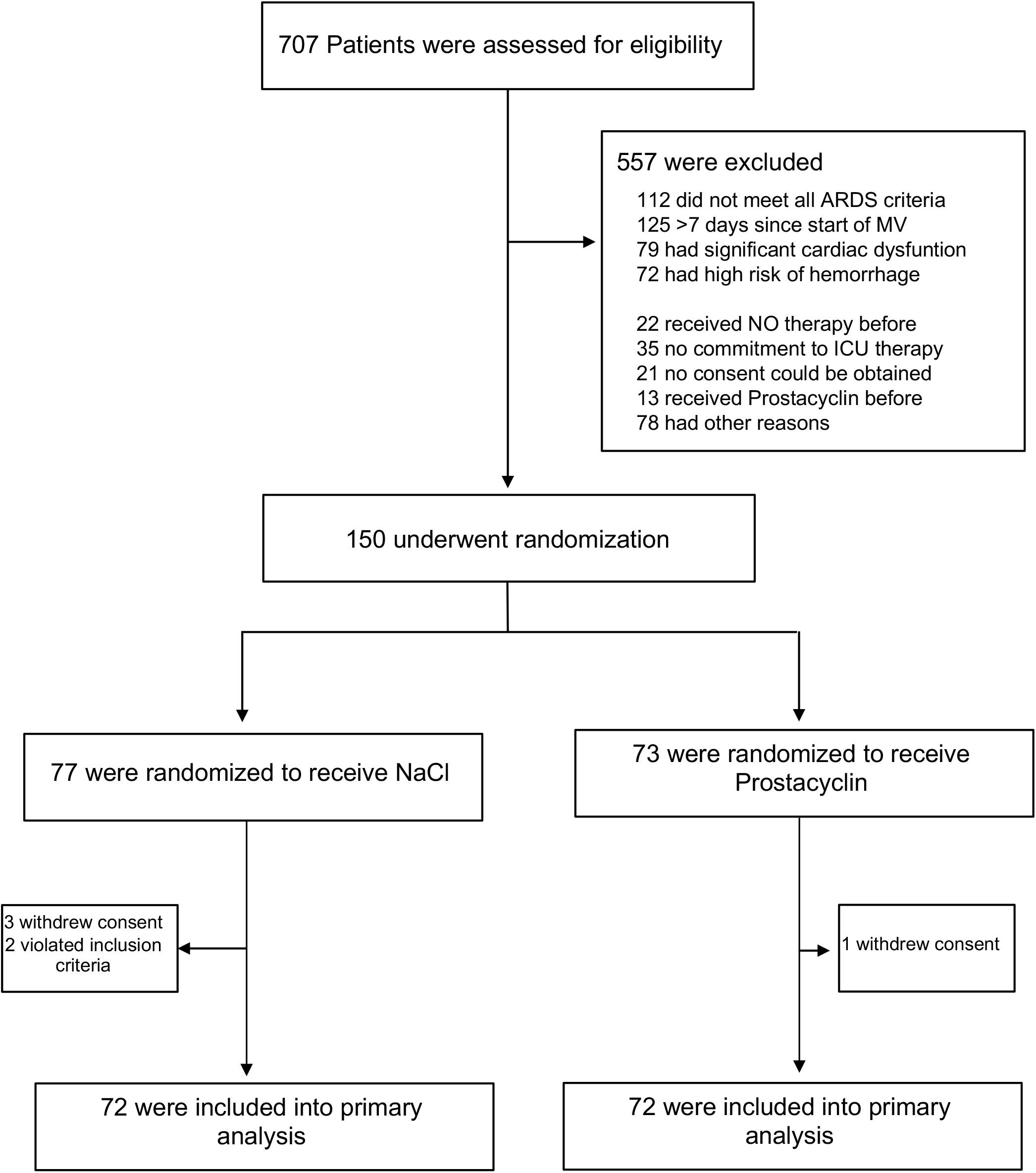
Screening, randomization, and follow-up of the study participants.

**Table 1:** Demographic and Baseline characteristics.

| Table 1: Demographic and Baseline characteristics |  |  |
| --- | --- | --- |
|  | Control (n=72) | Prostacyclin (n=72) |
| Age, mean ± SD, years | 56.0±14.0 | 61.1±14.4* |
| Weight, mean ±SD, kg | 93.6±20.7 | 93.3±23.8 |
| Height , mean ±SD, cm <sup>a</sup> | 174.4±9.2 | 174.4±9.2 |
| Body Mass Index <sup>a</sup> | 30.8±6.5 | 30.7±7.7 |
| Male | 55 (76%) | 53 (74%) |
| Female | 17 (24%) | 19 (26%) |
| **Causes of ARDS** |  |  |
| SARS-CoV2 | 52 (72%) | 49 (68%) |
| Aspiration | 3 (4%) | 4 (6%) |
| Viral Pneumonia (HSV etc.) | 2 (3%) | 1(1%) |
| Bacterial Pneumonia | 1 (1%) | 5 (7%) |
| Sepsis | 6 (8%) | 4 (6%) |
| Pancreatitis | 2 (3%) | 1 (1%) |
| Thoracic Trauma | 1 (1%) | 2 (3%) |
| Other | 5 (7%) | 6 (8%) |
| **Comorbidities, No. (%)** |  |  |
| Hypertension | 37 (51%) | 33 (46%) |
| unknown | 4 (6%) | 3 (4%) |
| Diabetes | 24 (33%) | 17 (24%) |
| COPD | 1 (1%) | 10 (14%)** |
| OSAS | 4 (6%) | 3 (4%) |
| Asthma | 5 (7%) | 2 (3%) |
| Sarcoidosis | 1 (1%) | 0 (0%) |
| Emphysema | 0 (0%) | 4 (6%)*** |
| Fibrosis | 0 (0%) | 1 (1%) |
| Tumor | 1 (1%) | 3 (4%) |
| LAE | 1 (1%) | 2 (3%)** |
| Chronic kidney disease (GFR<60) | 5 (7%) | 5 (7%) |
| Cardiac disease | 11 (15%) | 16 (22%) |
| Adipositas | 12 (17%) | 12 (17%) |
| Transplantation | 2 (3%) | 0 (0%) |
| HIV | 1 (1%) | 0 (0%) |
| Immune suppression | 5 (7%) | 2 (3%) |
| Psychiatric diseases | 4 (6%) | 12 (17%)* |
| Neurological diseases | 11 (15%) | 7 (10%) |
| Liver disease | 5 (7%) | 3 (4%) |
| Coagulopathy | 0 (0%) | 3 (4%) |
| Tumor (anamnestic) | 2 (3%) | 7 (10%) |
| OSAS | 4 (6%) | 3 (4%)** |
| SOFA Admission Score, mean ± SD <sup>b</sup> | 10.8 ± 3.2 | 10.8 ± 3.7 |
| **Reasons for ICU Admission** |  |  |
| Medical | 62 (86%) | 60 (83%) |
| Surgery | 4 (6%) | 2 (3%) |
| Emergency Surgery | 6 (8.3%) | 10 (14%) |

a= 142 patients included; b= 135 patients included
\*p=0.034, \*\*p=0.005, \*\*\*p=0.043

### Primary outcome

We defined the oxygenation index on Day 5 following treatment with the study drug as the primary outcome, and the oxygenation index at baseline was not significantly different between groups. Following treatment with prostacyclin, the oxygenation index showed a tendency to improve when considering all patients included in the trial.

Therefore, the primary group showed a strong tendency toward improvement (difference in improvement prostacyclin vs. NaCl groups of 19.5mmHg, baseline adjusted 20.1 mmHg, p=0·177, 95% CI (−9.1)-(+49·4)) following prostacyclin inhalation (Table 2, Figure 2). The interaction between the baseline and treatment arm was not significant (p=0·94). Sex (p=0·073, female vs. male 33·4 mmHg), age (0·11 mmHg per year, p=0·85), direct vs. indirect injury (indirect vs. direct injury 58·8mmHg, p=0·068), or COVID (no COVID vs. COVID 28.0 mmHg p=0·115) were not prognostic factors; however, differences might be relevant for each factor except for age (Supplemental Table 2, 3). When examining the subset of patients with COVID-19-induced ARDS, we observed a significant increase in the oxygenation index on Day 5 in patients treated with prostacyclin compared to patients with NaCl (34·4mmHg, p=0·043). The interaction between COVID-19 and treatment was not significant (p=0·104). For additional details, see Figure 2. Treatment effects were comparable for male patients (16·7 mmHg, p = 0·28) and the smaller subgroup of female patients (25.6mmHg, p = 0·49). A clear trend toward a larger treatment effect on elderly patients was observed, increasing from patients aged 20 to 39 years (−4·7mmHg, in favor of the control, p=0·85) to 24·4mmHg in patients aged 70 years or older (24·4 mmHg, p=0·45). However, the interaction between age and treatment was not significant (p=0·28). The effect on patients with direct injury was considerably larger (24·6mmHg, p=0·107) than that on the very small group of patients with indirect lung injury (−80·4 mmHg in favor of the control, p = 0·077). The interaction was significant (p=0·029).

**Table 2:** Main Clinical Outcomes.

| Table 2: Main Clinical Outcomes |  |  | p-value |
| --- | --- | --- | --- |
|  | Control (n=72) | Prostacyclin (n=72) |  |
| **Oxygenation Index** |  |  |  |
| Baseline | 123.6 ± 54.0<br>(111.0-136.2) | 123.2 ± 51.0<br>(11.3-135.0) | 0.96 |
| Day 5 | 208.6 ± 92.1<br>(186.9-230.4) | 227.9 ± 97.5<br>(204.7-251.1) | 0.24 |
| Difference Day 5 - Baseline<sup>a</sup> | 85.0 ± 84.3<br>(65.0-105.0) | 104.7 ± 90.5<br>(83.1-126.3) | 0.189* |
| Death at 90 days | 22 (31%, 20%-42%) | 23 (32%, (21%-44%)) |  |
| SOFA at day 7 <sup>c</sup> | 9.0 ± 4.7 (7.7-10.3) | 8.6 ± 4.7 (7.3-9.9) |  |
| SOFA at day 14 <sup>d</sup> | 9.7 ± 5.7 (7.7-11.8) | 10.5 ± 5.1 (8.7-12.3) |  |
| SOFA at day 28 <sup>e</sup> | 10.8 ± 5.7 (7.1-14.4) | 8.8 ± 5.6 (5.6-12.0) |  |
| Duration of ventilation |  |  |  |
| Including pauses in d | 11 (11-14, 8-14) | 11 (7-14, 9-14) |  |
| ICU length of Stay in d | 16 (10-34, 14-23) | 17 (12-43, 14-28)) |  |
| Ventilator Associated Pneumonia <sup>f</sup> | 5 (7%, 2%-15%) | 5 (7%, 2%-16%) |  |
| ICU Acquired Weakness <sup>g</sup> | 7 (10%, 4%-19%) | 4 (6%, 2%-14%) |  |
| **Discharge Location <sup>h</sup>** |  |  |  |
| Home | 20 (41%, 27%-58%) | 19 (40%, 26%-55%) |  |
| Skilled Nursing facility | 1 (2%, 0%-11%) | 1 (2%, >0%-11%) |  |
| Rehabilitation unit | 3 (6%, 1%-17%) | 6 (13%, 5%-25%) |  |
| Other transfer unit | 25 (51%, 36%-66%) | 22 (46%, 31%-61%) |  |

c= 109 patients included; d= 65 patients included; e= 26 patients included; f= 143 patients included; g= 140 patients included; h= 97 patients included \*p-value differs from baseline adjusted analysis (p=0.177), Entries are mean ± SD, median interquartile range or absolute and percentage frequency, results in brackets are 95% CIs for the mean or Interquartile ranges and 95% CIs for the median or 95% CIs for proportions. Death at 90 days RR = 1.05 (95% CI 0.93-1.18), Risk difference = 1.4% (95% CI (-13.8%) – (+16.5%).

**Figure 2.**
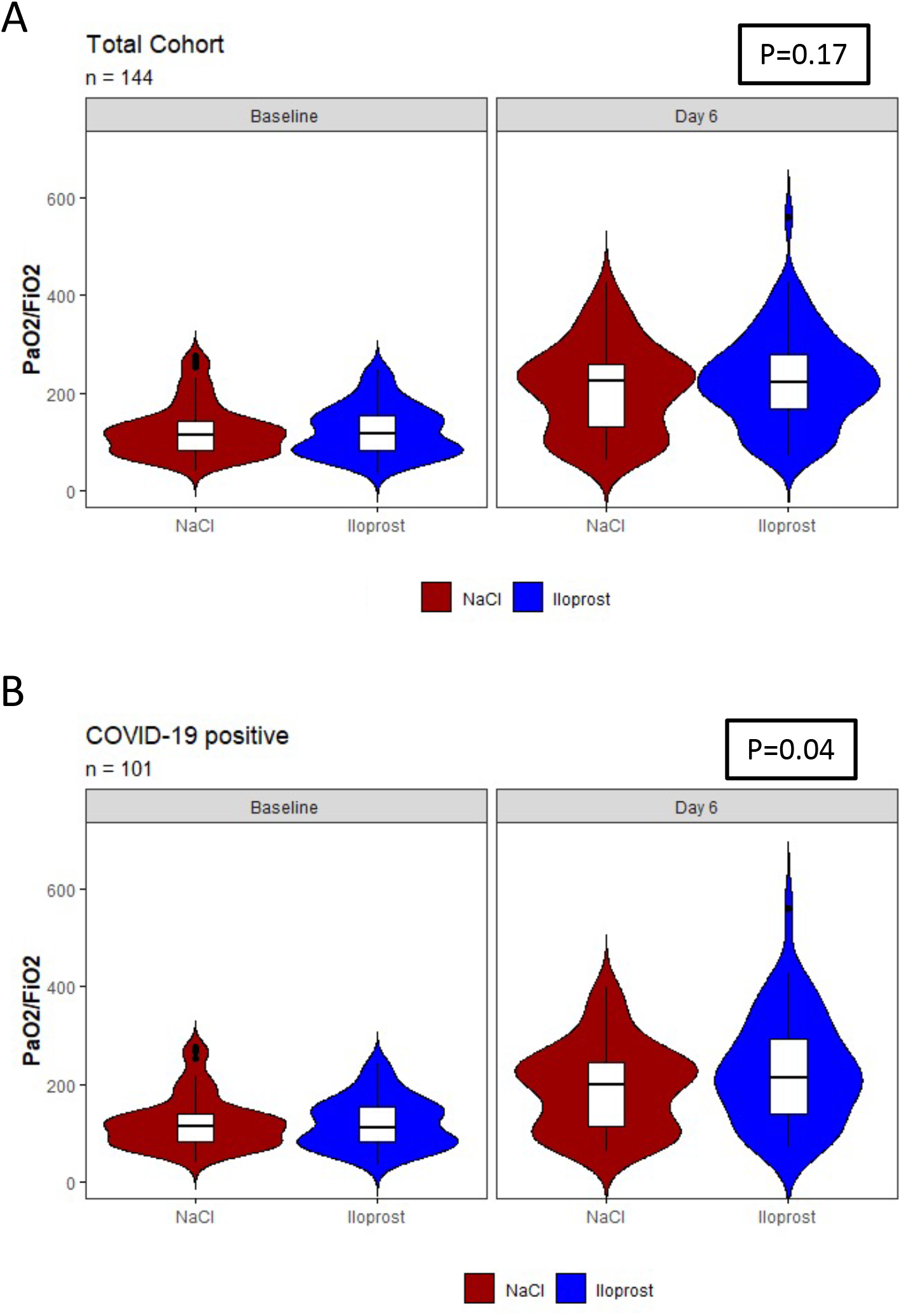
Oxygenation on Day 5 of treatment (day 6 following baseline) in the prostacyclin-treated group compared the control (NaCl)-treated group among **A)** all patients included in the trial and **B)** all COVID-19+ patients included in the trial.

### Secondary outcomes

Secondary outcomes were not significantly different between groups. Following treatment with prostacyclin, the mortality rate did not improve when analyzing all patients with ARDS (Figure 3). Regarding survival, no treatment differences were observed in any subgroup (p>0.4 in either male or female patients, in any age stratum, in patients with direct or indirect lung injury or in patients with or without COVID-19). In the total sample, no difference in the SOFA scores on Days 7, 14 and 28 were observed between study arms. The duration of mechanical ventilation and ICU length of stay did not differ between groups. The incidence of ventilator-associated pneumonia and ICU acquired weakness also did not differ between groups. The discharge location was also similar in both groups (Table 2).

**Figure 3.**
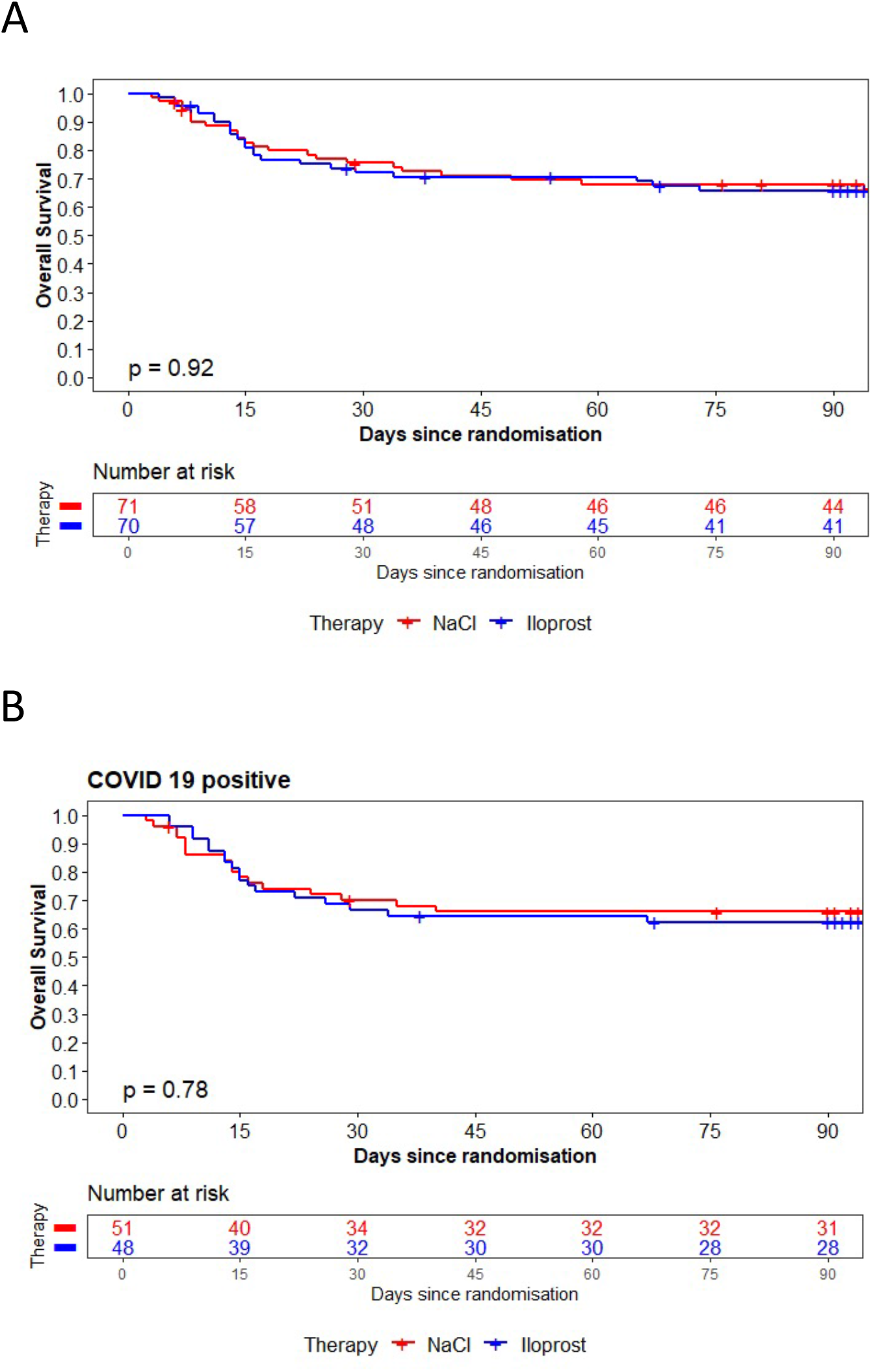
Ninety-day mortality rates in the prostacyclin-treated group compared with the control (NaCl)-treated group among A) all patients included in the trial and B) all COVID-19+ patients included in the trial.

When analyzing the subset of patients with COVID-19, we found that the secondary outcomes were not significantly different between groups. In this subgroup of patients, treatment with prostacyclin did not improve secondary outcomes. The SOFA score of patients with COVID-19 was not improved on Days 7, 14 and 28. The duration of mechanical ventilation and ICU length of stay did not differ between groups of patients with COVID-19. The incidence of ventilator-associated pneumonia, discharge location and ICU-acquired weakness also did not change in patients with COVID-19 following treatment with prostacyclin.

### Adverse events

Adverse events did not differ significantly between groups. In the treatment group, we identified a similar incidence of bleeding complications than in the NaCl group (9 vs. 11). Similar results were also obtained for the transfusion requirements. The incidence of thrombotic pulmonary embolism, coagulopathy, need for RRT and incidence of gastrointestinal complications also did not differ between groups. Neurological and cardiovascular complications were similar in both groups (Table 3).

**Table 3:** Adverse events.

| **Table 3: Adverse events** |  |  |
| --- | --- | --- |
|  | **Control** | **Prostacyclin** |
| Bleeding, No. (%) | 11 (15%, 8%-26%) | 9 (13%, 6%-22%) |
| Transfusion requirement (RBC), No. (%)<sup>i</sup> | 24 (34%, 23%-46%) | 24 (34%, 23%-46%) |
| Thrombotic Event, Pulmonary Embolism or Coagulopathy | 5 (7%, 2%-15%) | 5 (7%, 2%-15%) |
| Need for Renal Replacement Therapy | 17 (24%, 14%-35%) | 15 (21%, 12%-32%) |
| Gastrointestinal complications, No. (%) | 13 (18%, 10%-29%) | 7 (9%, 4%-19%) |
| Neurologic complications, No. (%) | 2 (3%, 0.3%-10%) | 4 (6%, 2%-14%) |
| Cardiovascular complications, No. (%) | 17 (24%, 14%-35%) | 13 (18%, 10%-29%) |

i= 142 patients included, results in brackets are 95% CIs for proportions

In patients with COVID-19, the incidence of adverse events was not significantly different between groups. We observed the same incidence of bleeding complications in the treatment group and the NaCl group. Similar results were obtained for the transfusion requirements. The incidence of thrombotic pulmonary embolism, coagulopathy, need for RRT and incidence of gastrointestinal complications also did not differ between groups. The incidences of neurological and cardiovascular were similar in both groups.

## Discussion

In this randomized controlled trial involving patients with ARDS, we addressed the question of whether inhaled prostacyclin would improve the lung function, as measured by oxygenation in the blood. We were able to show improved oxygenation on Day 6 of treatment in a population with ARDS however, the effect was not significant. The observed effect of prostacyclin was not associated with improved secondary outcomes in the intervention group, and neither the overall outcome nor the incidence of secondary complications was significantly different between groups.

In addition to extensive inflammation within the alveolar space, the central hallmark of ARDS is hypoxia (11, 12). Prone positioning and the use of extracorporeal membrane oxygenation (ECMO) have been shown to reduce hypoxia and to increase oxygenation (13, 14). ECMO therapy, however, is limited to expert centers and cannot be used widespread in all hospitals caring for these patients, since it involves a significant logistical effort and expert knowledge. Therefore, pharmaceutical approaches to improve pulmonary function are still very important. Several of these strategies have been tested previously without positive results. The use of aspirin in patients with ARDS did not result in a significant clinical improvement or better overall clinical outcome (4). The use of HMG-CoA reductase inhibitors was tested to improve overall outcomes and oxygenation in this patient population but did not exert a positive effect (5). Infusions of beta 2 agonists were also tested in patients with ARDS, but did not exert a positive effect on the outcome and oxygenation of patients with ARDS (15). The results described in this trial are the first to show that a prostacyclin intervention showed a tendency toward exerting a positive effect on oxygenation in critically ill patients with ARDS, especially in patients with COVID-19-induced ARDS. In a small case study of twenty patients, Sawheny et al. showed that oxygenation in patients with ARDS was improved by administering inhaled prostacyclin (9). However, this study was performed without a control group and did not employ a randomized prospective design. Therefore, no data from an RCT regarding the use of prostacyclin in patients with ARDS have been published to date.

As mentioned above, this randomized study is the first to document the effect of prostacyclin on patients with ARDS and COVID-19-induced ARDS. COVID-19-induced ARDS is an entity characterized by additional features compared to classical ARDS. Patients with COVID-19 present widespread pulmonary microthrombi and inflammatory infiltrates with diffuse pulmonary fibrosis (8, 16). In addition, endothelial dysfunction and a severe inflammatory response are indicators of COVID-19-induced pulmonary failure. Furthermore, hypoxemia that is unrelated to lung mechanics is present in patients with COVID-19-induced ARDS (17). These pathological features are patterns that could be improved by prostacyclin. Prostacyclin controls platelet aggregation and aggregability, preventing thrombus formation in an environment with a damaged endothelium (18, 19). In addition, prostacyclin interacts with and enhances the effect of nitric oxide on the vascular surface (20). As a result, endothelial function is improved, microthrombi are prevented, and the inflammatory response is reduced by administering prostacyclin to these patients. All of the described effects have important beneficial functions in patients with ARDS, especially in patients with COVID-19-induced ARDS, and might explain the positive effect we observed in this trial following the inhalation of prostacyclin.

Of course, our trial also has several limitations. First, the trial was started before the COVID-19 pandemic to evaluate the effects of prostacyclin on oxygenation and outcomes of critically ill patients with ARDS. Then, shortly after the start of the trial, the first wave of patients with COVID-19-induced ARDS were treated in Germany and German ICUs, including ours. Given the potential differences in the pathologies of ARDS and COVID-19-induced ARDS, this factor might have significant implications for therapy with prostacyclin. However, we decided to include all patient groups with ARDS and not exclude patients with COVID-19, since our trial should also take advantage of the opportunity to compare patients with different ARDS etiologies and their responses to prostacyclin treatment. Second, our sample size was moderate, and our study was probably underpowered. This interpretation seems justified, as we obtained the expected effect, i.e., a superiority of 21mmHg in PaO_2_/FiO_2_, but the standard deviations were much larger, as expected (80mmHg in the controls, 91mmHg in the prostacyclin group vs. 40 mmHg assumed). Third, the intervention group and the control group differed significantly in age, which could have a potential effect on the overall outcome in this patient group. The average age was older in the intervention group, and therefore, one would expect this factor to have a potential negative effect if any effect at all, based on the literature (21, 22). However, in our sample, no significant association of age with the primary outcome was observed. We also included patients receiving ECMO in this trial, which is particularly important because we measured oxygenation as the primary outcome. We recorded a nonsignificant difference between 21 patients treated with ECMO in the control group and 14 patients treated with ECMO in the treatment group, but of course, ECMO is important for the oxygenation levels measured. This is remarkable since the larger number in the control group would potentially skew the oxygenation toward the control group on Day 6, but we did not observe this result. The treatment groups still performed better when analyzing the primary outcome oxygenation and supported the positive effect of prostacyclin on oxygenation. Fourth, although the study medication assignment was randomized, we did not blind the investigators to the study medication, which was not possible due to the complex nature of the preparation of the prostacyclin in a blinded manner in our setting; therefore, we did not pursue this approach. Fifth, we included patients who had ARDS due to multiple reasons, and patients with and without COVID-19. However, impaired oxygenation is the common cardinal symptom of patients with all forms of ARDS, and most clinical approaches to improve oxygenation in all patients were tested in heterogeneous clinical ARDS groups, since we wanted to identify a commonly used intervention that would improve the poor oxygenation status. Therefore, we included all patients who met the inclusion criteria.

In conclusion, among patients with severe ARDS, inhaled prostacyclin showed a tendency to improve oxygenation. This change was not associated with a survival benefit but was associated with an improvement of secondary outcomes in the treated patient population. Larger clinical trials will evaluate the effect of prostacyclin on the overall outcomes of patients with ARDS.

## Data Availability

All data produced in the present study are available upon reasonable request to the authors

## Data sharing

After publication, the data will be made available to others on reasonable requests to the corresponding author. A proposal with detailed description of study objectives and statistical analysis plan will be needed for evaluation of the reasonability of requests. Additional materials might also be required during the process of evaluation. Data will be provided after approval from the University of Tübingen.

## Acknowledgments

We thank the study nurse team in Tübingen for their assistance with this project and all other personnel in the study centers for entering data into the eCRF file. We also thank all the study nurses at all participating centers for their work and thank the staff of the participating ICUs for their hard work.

## SUPPLEMENTAL DATA

## Methods

### Study design, Ethics and Oversight

A data safety and monitoring board oversaw the study and reviewed safety data periodically. Onsite monitoring for correctness of the consent procedure was performed at all sites by the Center for Clinical Studies Tübingen (Germany). The members of the writing committee wrote all drafts of the manuscript. All authors approved the final version of the manuscript and made the decision to submit it for publication.

### Patients inclusion and exclusion criteria

Patients received a study identification number and a treatment allocation at enrollment if inclusion criteria were met.

Inclusion criteria were: 1) PaO_2_/FiO_2_ ≤ 300 at time of ARDS diagnosis, 2) bilateral opacities on a frontal chest radiograph, 3) required positive pressure ventilation via an endotracheal tube or noninvasive ventilation, 4) no clinical signs of left atrial hypertension, and 5.) an “acute onset” defined as a duration of the hypoxemia criterion (#i) and the chest radiograph criterion (#ii) ≤ 28 days at the time of randomization. Patients were required to be randomized within 96 hours of the ARDS diagnosis and no later than 7 days from the initiation of mechanical ventilation.

Patients were excluded if: 1) the patient, surrogate or physician was not committed to full intensive care support, 2) patients had a positive pregnancy test at the time of screening, 3) had contraindications for the use of prostacyclin 4) received nitric oxide or prostacyclin therapy within the previous 24 h before study randomization, or 5) were dependent on the sponsor, investigator, manufacturer and/or their employees.

### Sample size

In our study, we assumed an effect size of 0.525, leading to 116 error degrees of freedom to achieve a power of 80% using a level of significance of 0.05 in the two-sided t-test. The interindividual effect size of 0.525 combined with the standard deviation of 40 corresponds to a difference of approximately 21 in the p_a_O_2_/FiO_2_ratio in treatment compared to controls. Therefore, we calculated numbers of patients to be assessed for eligibility (n=300), assigned to the trial (n = 150), and analyzed (n=150 in the intention-to-treat analysis). The sample size and power consideration referred to 120 evaluable patients.

### Statistical analysis

Additionally, the interaction between baseline and treatment was tested and included in the model if the result was significant. In the case of an interaction, the main effect was retrieved for the arithmetic mean of baseline values using the centered variable for p_a_O_2_/FiO_2_. Multiple imputation was applied. The level of significance was 0.05 (two-sided), and no interim analysis was performed. Only the primary analysis was confirmatory. Subgroup analyses were planned for sex and race, patients with increased pulmonary arterial pressure, direct or indirect lung injury, and age by decades, but only in groups with 40 patients or more. In the final analysis, we combined age groups, and we also report results from strata of sizes slightly smaller than 40. An additional primarily unplanned subgroup analysis was performed for patients with COVID, as the pandemic only occurred during the course of the study. The same factors were also tested for prognostic value.

The statistical analysis of the prespecified secondary endpoints was performed with descriptive and exploratory statistical methods according to the scale and observed distribution. P values were reported, although all secondary analyses are nonconfirmatory.

## SUPPLEMENTAL TABLES

**Supplemental Table 1:** Organ Specific Baseline Characteristics and Ventilation Parameter.

| **Supplemental Table 1: Organ Specific Baseline Characteristics and Ventilation Parameters** |  |  |
| --- | --- | --- |
|  | **Control (n=72)** | **Prostacyclin (n=72)** |
| ECMO yes/ no <sup>i</sup> | 21 (29%) | 15 (21%)<sup>m</sup> |
| COVID-19 + ECMO | 17 (23%) | 14 (19%) |
| Duration of ECMO (days), median (IQR) | 17 (11-38) | 27 (9-55) |
| Inspiratory Plateau Pressure cmH<sub>2</sub>O median (IQR) | 20 (18-23) | 20 (17-23) |
| Tidal Volume/ kg predicted body weight | 6.3 ± 2.5 | 6.7 ± 1.9 |
| Driving Pressure cmH<sub>2</sub>O | 12 (9-15) | 13 (11-16) |
| Acidosis | 24 (34%) | 36 (50%)* |
| Lactate level (Minimum), Median (IQR) mmol/l | 1.00 (0.80-1.20) | 0.95 (0.70-1.38) |
| Lactate level (Maximum) , Median (IQR) mmol/l | 1.40 (1.13-1.80) | 1.5 (1.10-2.18) |
| Alanine Aminotransferase (U/l) ALT <sup>j</sup> , median (IQR) | 34 (22-54) | 35 (24-68) |
| Aspartate Aminotransferase (U/l) AST <sup>k</sup> | 48 (32-64) | 59 (37-96) |
| INR | 1.2 (1.1-1.6) | 1.2 (1.1-1.5) |
| Bilirubin (Median, IQR) | 0.7 (0.4-1.2) | 0.6 (0.4-1.1) |
| Creatine Kinase (U/l) | 287 (112-768) | 261 (75-730) |
| BUN, median (IQR) <sup>l</sup> | 46 (30-64) | 52 (35-68) |
| Hemoglobin level (g/l) | 10.0 ± 2.3 | 10.4 ± 2.2 |
| Platelet Count (nx10<sup>3</sup>/μl) | 231 (167-300) | 207 (144-307) |
| Vasopressor Dependent No | 62 (87%) | 61 (85%) |
| Lowest Mean Arterial Blood Pressure mmHg | 65.1 ± 8.9 | 64.2 ± 7.6 |

Entries are mean ± SD, results in brackets are 95% CIs for the mean, i= 142 patients included; j= 119 patients included; k= 117 patients included; l= 143 patients included<sup>m</sup>p = 0.05

**Supplemental Table 2:** Primary Endpoint, COVID 19 patients only (imputated data, n=101 each analysis)

| **Oxygenation Index** | **Control** | **Prostacyclin** | **p-value** |
| --- | --- | --- | --- |
| Baseline | 121.6 ± 53.3 (107.0-136.3) | 121.0 ± 50.6 (106.6-135.3) | 0.95 |
| Day 5 | 193.5 ± 88.4 (168.9-218.2) | 227.3 ± 109.0 (196.1-258.4) | 0.093 |
| Difference Day 5 - Baseline<sup>a</sup> | 71.9 ± 78.0 (50.0-93.7) | 106.3 ± 96.5 (78.7-134.0) | 0.054* |

Entries are mean ± SD, results in brackets are 95% CIs for the mean, \*p value differs from baseline adjusted analysis (p=0.043)

**Supplemental Table 3:** Primary Endpoint in age strata (imputed data, n=144 in each analysis)

| **Oxygenation Index** | **Control** | **Prostacyclin** |
| --- | --- | --- |
| **20-<40 ys** |  |  |
| Baseline | 123.9 ± 73.5 | 121.7 ± 49.6 |
| Day 5 | 248.0 ± 135.4 | 245.2 ± 97.5 |
| Difference Day 5 - Baseline | 124.1 ± 109.4 | 123.5 ± 98.1 |
| **40-<60 ys** |  |  |
| Baseline | 110.2 ± 46.8 | 113.6 ± 123.3 |
| Day 5 | 191.8 ± 92.4 | 211.6 ± 53.1 |
| Difference Day 5 - Baseline | 81.5 ± 86.5 | 98.0 ± 109.4 |
| **60-<70 ys** |  |  |
| Baseline | 133.4 ± 48.3 | 120.0 ± 43.5 |
| Day 5 | 211.4 ± 67.8 | 221.2 ± 87.2 |
| Difference Day 5 - Baseline | 78.0 ± 66.5 | 101.2 ± 77.9 |
| **70ys and older** |  |  |
| Baseline | 154.5 ± 52.8 | 137.8 ± 53.7 |
| Day 5 | 229.0 ± 63.1 | 247.4 ± 68.0 |
| Difference Day 5 - Baseline | 74.5 ± 71.0 | 109.6 ± 76.1 |

Entries are mean ± SD, 20-<40 ys n=16, 40-<60 ys n=56, 60-<70 ys n=44, 70+ ys n=38 All p-values were larger than 0.4

**Supplemental Figure 1.**
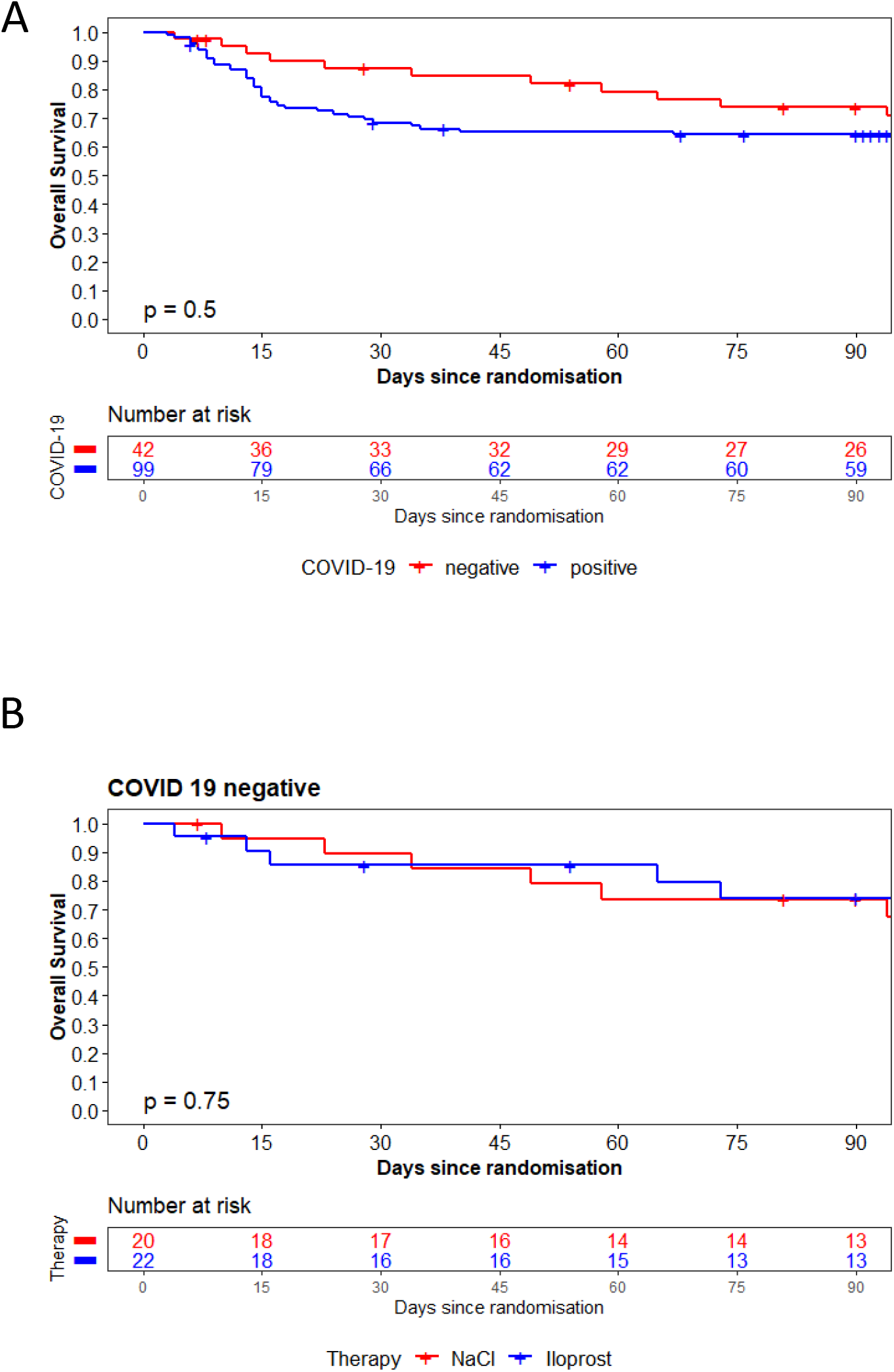
A) Ninety-day mortality rates in COVID-19-negative and COVID-19-positive patients and B) 90-day mortality rates in COVID-19-negative patients in the prostacyclin-treated group compared with control (NaCl)-treated patients.

